# TRENDS IN CASES, HOSPITALISATION AND MORTALITY RELATED TO THE OMICRON BA.4/BA.5 SUB-VARIANTS IN SOUTH AFRICA

**DOI:** 10.1101/2022.08.24.22279197

**Authors:** Waasila Jassat, Salim S Abdool Karim, Lovelyn Ozougwu, Richard Welch, Caroline Mudara, Maureen Masha, Petro Rousseau, Milani Wolmarans, Anthony Selikow, Nevashan Govender, Sibongile Walaza, Anne von Gottberg, Nicole Wolter, Pedro Terrence Pisa, Ian Sanne, Sharlene Govender, DATCOV author group, Lucille Blumberg, Cheryl Cohen, Michelle J. Groome

## Abstract

**Introduction:** The Omicron BA.1/BA.2 wave in South Africa had lower hospitalisation and mortality than previous SARS-CoV-2 variants and was followed by an Omicron BA.4/BA.5 wave. This study compared admission incidence risk across waves, and the risk of mortality in the Omicron BA.4/BA.5 wave, to the Omicron BA.1/BA.2 and Delta waves.

**Methods:** Data from South Africa’s national hospital surveillance system, SARS-CoV-2 case linelist and Electronic Vaccine Data System were linked and analysed. Wave periods were defined when the country passed a weekly incidence of 30 cases/100,000 people. Mortality rates in the Delta, Omicron BA.1/BA.2 and Omicron BA.4/BA.5 wave periods were compared by post-imputation random effect multivariable logistic regression models.

**Results:** In-hospital deaths declined 6-fold from 37,537 in the Delta wave to 6,074 in the Omicron BA.1/BA.2 wave and a further 7-fold to 837 in the Omicron BA.4/BA.5 wave. The case fatality ratio (CFR) was 25.9% (N=144,798), 10.9% (N=55,966) and 7.1% (N=11,860) in the Delta, Omicron BA.1/BA.2, and Omicron BA.4/BA.5 waves respectively. After adjusting for age, sex, race, comorbidities, health sector and province, compared to the Omicron BA.4/BA.5 wave, patients had higher risk of mortality in the Omicron BA.1/BA.2 wave (adjusted odds ratio [aOR] 1.43; 95% confidence interval [CI] 1.32-1.56) and Delta (aOR 3.22; 95% CI 2.98-3.49) wave. Being partially vaccinated (aOR 0.89, CI 0.86-0.93), fully vaccinated (aOR 0.63, CI 0.60-0.66) and boosted (aOR 0.31, CI 0.24-0.41); and prior laboratory-confirmed infection (aOR 0.38, CI 0.35-0.42) were associated with reduced risks of mortality.

**Conclusion:** Overall, admission incidence risk and in-hospital mortality, which had increased progressively in South Africa’s first three waves, decreased in the fourth Omicron BA.1/BA.2 wave and declined even further in the fifth Omicron BA.4/BA.5 wave. Mortality risk was lower in those with natural infection and vaccination, declining further as the number of vaccine doses increased.

## INTRODUCTION

Until the emergence of the Omicron variant in November 2021, each new variant of concern that spread in South Africa led to more infections, hospitalisations and deaths in South Africa. The Omicron variant of concern (VOC) marked a shift in the COVID-19 pandemic. The Omicron BA.1 sub-variant, while more transmissible and associated with immune escape, was reported to be less virulent as it showed attenuated replication in mice (1) and infected upper more than lower respiratory tract (2). It emerged at a time when 73% of South Africans had developed hybrid immunity through vaccination and/or prior infection (3). As a result of high levels of immunity and a less virulent variant, in the fourth Omicron BA.1 dominated wave in South Africa, lower levels of hospitalisation, severity and mortality were observed compared to previous waves dominated by D614G, Beta and Delta variants (4,5). Reduced severity with Omicron BA.1 was also reported in other countries (6-9). However, a study in Hong Kong reported similar mortality among individuals infected with Omicron who did not have pre-existing immunity, to those infected with earlier variants (10).

After the Omicron BA.1/BA.2 wave, a number of additional Omicron sub-variants (rather than a new variant of concern) caused resurgences in many parts of the world. In South Africa, BA.2 was responsible for sustained transmission in late January 2022 at the tail-end of the BA.1 surge, while BA.3 never became dominant and circulated at low levels (11). Omicron BA.4 and BA.5 were detected in South Africa in February 2022 (12) and both jointly dominated the fifth wave from April-June 2022. Shortly after having experienced waves related to Omicron BA.1/BA.2, Omicron BA.4 and/or BA.5 became dominant globally and cases increased in many countries (13).

Omicron sub-variants are more transmissible than previous VOCs, with greater immune escape, with BA.4/BA.5 showing reduced neutralization from antibodies induced by BA.1 infection, more so in the unvaccinated (14,15). Early data showed that Omicron BA.4/BA.5 waves had reduced severity in South Africa (16) and the United States (17).

It remains important to understand the characteristics of severe disease caused by new VOCs or viral lineages in different geographic locations in order to guide public health policy and planning. The purpose of this study was to investigate trends in the incidence of laboratory-confirmed COVID-19 hospitalisations by age group in each COVID-19 wave in South Africa and compare the risk of mortality in the most recent Omicron BA.4/BA.5 wave with the Omicron BA.1 and the Delta waves.

## METHODS

Data on real-time reverse transcription polymerase chain reaction (rRT-PCR) and antigen positive SARS-CoV-2 cases were collated daily from laboratory reports while data on COVID-19 hospital admissions were collected through DATCOV, an active surveillance programme established specifically for COVID-19. These case and hospital surveillance systems have been previously described (18).

Data for the period from the first case in South Africa on 5 March 2020 to 28 May 2022 were analysed. Three data sources were linked for this analysis using South African identification numbers, first names, surnames and dates of birth, (1) the DATCOV national COVID-19 hospital surveillance established by the National Institute for Communicable Diseases, (2) the Electronic Vaccine Data System (EVDS) established by the National Department of Health, and (3) the national SARS-CoV-2 case linelist, the Notifiable Medical Conditions Surveillance System (NMC-SS).

Incidence risks were calculated using Statistics South Africa mid-year population figures for 2021 (19). A weekly incidence of 30 cases/100,000 population was the threshold for the start and end of each wave, thereby defining the distinct periods of analysis, namely D614G (first) wave from 7 June – 22 August 2020, Beta (second) wave from 15 November 2020 – 6 February 2021, Delta (third) wave from 9 May – 18 September 2021, Omicron BA.1/BA.2 (fourth) wave from 28 November 2021 – 5 February 2022, and Omicron BA.4/BA.5 (fifth) wave from 17 April – 28 May 2022.

Analysis of mortality was restricted to admissions that had already accumulated outcomes and all patients still in-hospital or transferred to other hospitals without final outcomes were excluded, because they remained at risk of still developing severe outcomes including death. Descriptive statistics were used to describe the trends in numbers of cases, admissions, mortality and case fatality ratios (CFR) over the equivalent periods of the D614G, Beta, Delta, Omicron BA.1/BA.2, and Omicron BA.4/BA.5 waves.

Post-imputation random effect (on admission facility) multivariable logistic regression models were used to compare mortality between the Delta, Omicron BA.1/BA.2 and Omicron BA.4/BA.5 waves. To account for incomplete or missing data on selected variables, we used multivariate imputation by chained equation (MICE) and generated ten complete imputed datasets that were used for subsequent analyses. Variables analysed using MICE included race and comorbidities, where up to a third of the data were missing. Complete or near-complete variables included in the imputation process were age, sex, province, health sector (i.e., public or private), in-hospital outcome (i.e., discharged alive or died), and vaccination status (See supplementary table 1 for variable completeness).

Age, sex, race, presence of a comorbidity (which included hypertension, diabetes, chronic cardiac disease, chronic kidney disease, asthma/chronic pulmonary disease, malignancy, HIV or tuberculosis), type of health sector (private or public), province, vaccination status and recorded laboratory-confirmed prior infection were included in the model to assess the relationship between each wave period and mortality in SARS-CoV-2 positive patients admitted to hospital. Prior infection was determined through linking to the NMC-SS and regarded as affirmative if an individual had a recorded positive SARS-CoV-2 test more than 90 days after a previous positive test. Vaccination status was determined at date of admission through EVDS linkage and individuals were considered to be unvaccinated if they had not received any COVID-19 vaccine doses, partially vaccinated if they received one dose of BNT162b, fully vaccinated if they received two doses of BNT162b or one dose of Ad26.COV2.S with the most recent dose at least 14 days earlier, and boosted if they received at least one additional COVID-19 vaccine dose of any kind in addition to full vaccination. [South Africa first rolled out COVID-19 vaccination among health care workers in February 2021, using Ad26.COV2.S under the Sisonke programme. In May 2021, vaccination using BNT162b or Ad26.COV2.S was introduced for individuals older than 60 years, then expanding to those aged 35-50 years in July 2021, 18-35 years in August 2021, and 12-18 years in October 2021. In December 2021, booster doses were introduced.]

The statistical analysis was implemented using Stata 15 (Stata Corp®, College Station, Texas, USA). We followed STROBE guideline recommendations. Ethical approval was obtained from the Human Research Ethics Committee (Medical) of University of the Witwatersrand for the collection of COVID-19 case data (M210752), and for the DATCOV surveillance programme (M2010108).

## RESULTS

The 7-day moving average of daily cases had a peak that was higher in each successive peak of the first four waves but lower in the fifth wave (Figure 1). The number of hospital admissions in each of the five waves were 78,529 (D614G), 118,160 (Beta), 163,887 (Delta), 61,942 (Omicron BA.1/BA.2) and 16,447 (Omicron BA.4/BA.5). Unlike the pattern observed in the prior waves, the rise in cases during the Omicron BA.1/BA.2 and Omicron BA.4/BA.5 waves were not accompanied by a concomitant rise in hospital admissions and in-hospital deaths.

**Figure 1:**
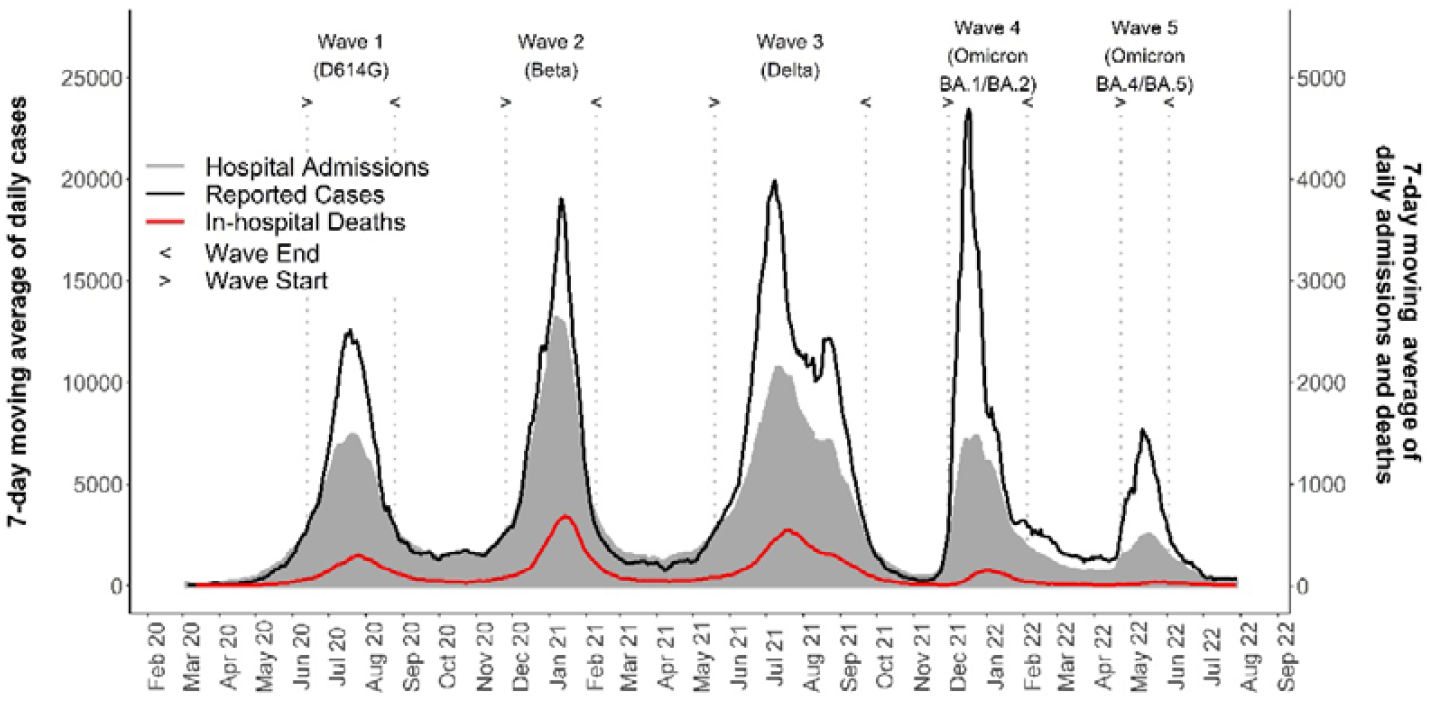
7 day moving average of SARS-CoV-2 cases, COVID-19 admissions and in-hospital deaths in South Africa, 5 March 2020-28 May 2022.

The incidence of COVID-19 admissions per 100,000 population was highest in ≥60 years in each wave (Table 1 and Supplementary Figure S1). In the fourth and fifth waves, incidence was highest in ≥60 years followed by <1 years. In adults and children aged 5-18 years, admission incidence risk increased from the first to the third wave then decreased in the fourth and fifth waves to the lowest incidence seen in any previous waves. In children aged ≤5 years, admission incidence risk increased from the first to the fourth wave, then decreased in the fifth wave to similar incidence as the second wave.

**Table 1:** COVID-19 admissions incidence risk per 100,000 by age group (in years) and wave period, South Africa, 5 March 2020-28 May 2022 (N=438,965)

| Age group (population size) | Incidence risk per 100,000 population, by wave period |  |  |  |  |
| --- | --- | --- | --- | --- | --- |
|  | Incidence risk (number of admissions) |  |  |  |  |
|  | Wave 1 D614G | Wave 2 Beta | Wave 3 Delta | Wave 4 Omicron BA.1/BA.2 | Wave 5 Omicron BA.4/BA.5 |
| <1 year (1,175,632) | 41.4 (487) | 80.7 (949) | 167.8 (1,973) | 184.4 (2,168) | 83.0 (976) |
| 1-4 years (4,533,324) | 8.8 (401) | 17.2 (779) | 37.5 (1,700) | 46.3 (2,100) | 19.3 (877) |
| 5-18 years (15,299,104) | 9.5 (1,458) | 12.1 (1,848) | 33.7 (5,154) | 27.7 (4,240) | 8.3 (1,270) |
| 19-39 years (21,624,489) | 77.6 (16,780) | 96.6 (20,879) | 149.0 (32,223) | 96.7 (20,920) | 17.8 (3,842) |
| 40-59 years (12,005,082) | 271.5 (32,591) | 385.7 (46,302) | 506.3 (60,778) | 125.2 (15,031) | 29.4 (3,532) |
| 60+ years (5,505,347) | 487.9 (26,863) | 860.4 (47,367) | 1128.6 (62,131) | 337.1 (18,560) | 99.6 (5,482) |
| Total (60,142,978) | 130.7 (78,580) | 196.4 (118,124) | 272.6 (163,959) | 104.8 (63,019) | 26.6 (15,979) |

The in-hospital CFR was 25.9% (37,537/144,798), 10.9% (6,074/55,966) and 7.1% (837/11,860) respectively in the Delta, Omicron BA.1/BA.2, and Omicron BA.4/BA.5 periods respectively (Table 2). The age specific CFRs for each wave show that the CFR was higher in older age groups across all waves; and was lowest in each age group in the Omicron BA.1/BA.2 and Omicron BA.4/BA.5 waves (Table2).

**Table 2:** COVID-19 in-hospital case fatality ratio (CFR) by age group (in years) and wave period, South Africa, 5 March 2020-28 May 2022 (N=438,965)

| Age group | Case fatality ratio (CFR) by wave period |  |  |  |  |
| --- | --- | --- | --- | --- | --- |
|  | CFR % (number of deaths/total admissions with outcomes) |  |  |  |  |
|  | Wave 1 D614G | Wave 2 Beta | Wave 3 Delta | Wave 4 Omicron BA.1/BA.2 | Wave 5 Omicron BA.4/BA.5 |
| <1 year | 6.7 (37/552) | 5.3 (44/835) | 6.1 (107/1,766) | 2.5 (50/1,986) | 1.3 (10/779) |
| 1-4 years | 0.9 (4/466) | 3.1 (21/675) | 1.3 (20/1,550) | 1.3 (25/1,947) | 0.3 (2/755) |
| 5-18 years | 3.7 (57/1,528) | 3.7 (61/1,638) | 2.9 (131/4,560) | 1.6 (60/3,869) | 1.1 (12/1,074) |
| 19-39 years | 6.7 (1,207/17,896) | 9.8 (1,811/18,403) | 9.0 (2,549/28,427) | 4.8 (887/18,685) | 3.9 (110/2,842) |
| 40-59 years | 17.2 (5,609/32,683) | 22.8 (9,286/40,789) | 22.9 (12,282/53,701) | 11.4 (1,515/13,260) | 6.8 (175/2,558) |
| 60+ years | 37.4 (9,916/26,536) | 43.5 (18,148/41,703) | 41.0 (22,448/54,794) | 21.8 (3,537/16,219) | 13.7 (528/3,852) |
| Total | 21.1 (16,630/79,661) | 28.2 (29,371/104,043) | 25.9 (37,537/144,798) | 10.9 (6,074/55,966) | 7.1 (837/11,860) |

Vaccination coverage amongst 144,798 patients admitted in the Delta wave was 12,747 (8.8%) partially vaccinated, 5,481 (3.8%) fully vaccinated, and 2 (0%) boosted, while 126,568 (87.4%) were unvaccinated. Amongst 55,966 patients admitted in the Omicron BA.1/BA.2 wave, 1,967 (3.5%) were partially vaccinated, 13,402 (23.9%) were fully vaccinated, and 433 (0.7%) were boosted, while 40,164 (71.8%) were unvaccinated. Amongst 11,860 patients admitted in the Omicron BA.4/BA.5 wave, 365 (3.1%) were partially vaccinated, 3,152 (26.6%) were fully vaccinated, and 1,016 (8.6%) were boosted, while 7,329 (61.8%) were unvaccinated.

On multivariable analysis (Table 3), after adjusting for age, sex, race, presence of a comorbidity, health sector (private/public) and province, compared to the Omicron BA.4/BA.5 wave, patients had a higher risk of mortality in the Omicron BA.1 (adjusted odds ratio [aOR] 1.43; 95% confidence interval [CI] 1.32-1.56) and Delta (aOR 3.22; 95% CI 2.98-3.49) wave periods. Being partially vaccinated (aOR 0.89, CI 0.86-0.93), fully vaccinated (aOR 0.63, CI 0.60-0.66) and boosted (aOR 0.31, CI 0.24-0.41); and prior laboratory-confirmed infection (aOR 0.38, CI 0.35-0.42) were associated with reduced mortality (Table 3).

**Table 3.** Factors associated with mortality among SARS-CoV-2 positive hospitalised patients in the Delta (9 May-18 Sep 2021), Omicron BA.1/BA.2 (28 November 2021-5 February 2022) and Omicron BA.4/BA.5 waves (17 April-28 May 2022), South Africa. (univariate and multivariable analysis implemented on the imputed dataset) (N=212,624)*

| Characteristic | Unimputed CFR<br>% (n/N) | Unadjusted OR<br>(95% CI) | Adjusted OR<br>(95% CI) | p value |
| --- | --- | --- | --- | --- |
| **Age group (years)** |  |  |  |  |
| <20 | 2.3 (447/19,533) | Reference | Reference |  |
| 20-39 | 7.2 (3,516/48,707) | 3.2 (2.9-3.5) | 2.8 (2.5-3.1) | <0.001 |
| 40-59 | 20.1 (13,972/69,519) | 10.7 (9.7-11.8) | 7.7 (7.0-8.5) | <0.001 |
| 60+ | 35.4 (26,513/74,865) | 23.5 (21.3-25.9) | 17.9 (16.2-19.8) | <0.001 |
| **Sex** |  |  |  |  |
| Female | 19.2 (22,410/116,800) | Reference | Reference |  |
| Male | 23.0 (22,018/116,800) | 1.3 (1.3-1.3) | 1.3 (1.3-1.3) | <0.001 |
| **Race** |  |  |  |  |
| White | 24.2 (4,441/18,386) | Reference | Reference |  |
| Mixed | 25.9 (2,069/7,976) | 0.8 (0.8-0.9) | 1.1 (1.1-1.2) | 0.0017 |
| Black | 21.5 (19,694/91,675) | 0.6 (0.6-0.6) | 1.1 (1.1-1.2) | 0.0003 |
| Indian | 23.0 (1,285/5,580) | 1.0 (0.9-0.9) | 1.2 (1.2-1.4) | <0.001 |
| Other | 21.7 (97/448) | 0.7 (0.5-0.8) | 0.9 (0.7-1.2) | 0.62 |
| **Comorbid condition\*\*** |  |  |  |  |
| No comorbidity | 14.5 (1,866/12,846) | Reference | Reference |  |
| Comorbid condition | 31.0 (2,799/9,025) | 2.4 (2.3-2.4) | 1.5 (1.4-1.5) | <0.001 |
| **Health sector** |  |  |  |  |
| Private sector | 17.4 (18,105/103,916) | Reference | Reference |  |
| Public sector | 24.2 (26,343/108,708) | 1.6 (1.4-1.9) | 1.4 (1.2-1.6) | <0.001 |
| **Province** |  |  |  |  |
| Western Cape | 20.3 (8,160/40,250) | Reference | Reference |  |
| Eastern Cape | 25.0 (3,616/14,492) | 1.3 (1.0-1.7) | 1.3 (1.1-1.6) | 0.0027 |
| Free State | 20.6 (2,656/12,871) | 1.1 (0.8-1.5) | 1.1 (0.8-1.3) | 0.65 |
| Gauteng | 21.1 (15,513/73,627) | 1.0 (0.8-1.2) | 1.2 (1.0-1.5) | 0.26 |
| KwaZulu-Natal | 18.6 (5,951/32,079) | 0.9 (0.7-1.1) | 1.1 (1.0-1.4) | 0.15 |
| Limpopo | 25.1 (2,460/9,805) | 1.8 (1.3-2.4) | 1.6 (1.2-1.9) | 0.0001 |
| Mpumalanga | 21.7 (2,184/10,081) | 1.6 (1.2-2.2) | 1.4 (1.1-1.8) | 0.0075 |
| North West | 18.3 (2,738/14,967) | 0.8 (0.6-1.2) | 0.8 (0.6-1.1) | 0.15 |
| Northern Cape | 26.3 (1,170/4,452) | 1.8 (1.2-2.7) | 1.5 (1.1-2.0) | 0.014 |
| **Wave period** |  |  |  |  |
| Omicron BA.4/BA.5 | 7.1 (837/11,860) | Reference | Reference |  |
| Delta | 25.9 (37,537/144,798) | 4.5 (4.2-4.9) | 3.2 (3.0-3.5) | <0.001 |
| Omicron BA.1/BA.2 | 10.9 (6,074/55,966) | 1.5 (1.4-1.6) | 1.4 (1.3-1.6) | <0.001 |
| **Prior infection** |  |  |  |  |
| No | 21.5 (43,906/204,326) | Reference | Reference |  |
| Yes | 6.5 (542/8,298) | 0.3 (0.3-0.3) | 0.4 (0.4-0.4) | <0.001 |
| **Vaccination** |  |  |  |  |
| No | 21.5 (37,421/174,061) | Reference | Reference |  |
| Partial | 28.9 (4,355/15,077) | 1.6 (1.5-1.7) | 0.9 (0.9-0.9) | <0.001 |
| Full | 5.6 (2,609/22,035) | 0.6 (0.5-0.6) | 0.6 (0.6-0.7) | <0.001 |
| Boosted | 4.3 (63/1,451) | 0.2 (0.2-0.3) | 0.3 (0.2-0.4) | <0.001 |

CFR: case fatality ratio; OR: Odds Ratio; CI: Confidence Interval
\* random effects multivariate logistic regression model controlling for clustering by facility
\*\* comorbid conditions included hypertension, diabetes, chronic cardiac disease, chronic kidney disease, asthma/chronic pulmonary disease, malignancy, HIV and active and past TB

## DISCUSSION

The fifth COVID-19 wave in South Africa, due predominantly to Omicron BA.4 and BA.5 sub-variants had a lower peak number of cases, risk of hospital admission and risk of in-hospital mortality compared to all the previous waves. Vaccination and prior infection were protective against COVID-19 mortality.

The reduced severity in the Omicron BA.1/BA.2 wave was thought to be related to multiple factors including a less virulent variant and widespread immunity due to vaccination and natural infection in South Africa (3-5). A combination of natural infection and vaccination, referred to as hybrid immunity, has been shown to protect better against Omicron BA.1 symptomatic COVID-19 than infection-only immunity (20).

While some South African studies have suggested that Omicron BA.4 and BA.5 had similar disease severity to Omicron BA.1 and BA.2 (16,21), our data indicate a lower risk of mortality in the Omicron BA.4/BA.5 wave compared to the Omicron BA.1/BA.2 wave. Immunological observations from South Africa point to considerable immune escape of Omicron BA.4 and BA.5 from Omicron BA.1 elicited immunity, but much less so in those with vaccination (15). In other settings such as the United States, a higher risk of breakthrough infection with Omicron BA.4 or BA.5 was observed among previously vaccinated or infected individuals, indicating greater immune escape, but these breakthrough infections were associated with low severity (17). Cell-mediated immunity likely contributed to the lower levels of hospitalisation and mortality in the Omicron BA.4/BA.5 wave (22).

A South African study with prospective ascertainment of repeat infections demonstrated that prior infection whether symptomatic or not, provided durable protection against reinfection beyond a year and resulted in a complex immune landscape (23). In contrast, Hong Kong’s Omicron BA.2.2 outbreak and high mortality among older individuals, could be explained by low immunity (the incidence of COVID-19 was very low in Hong Kong as a result of pandemic control efforts and protection for most individuals was likely elicited by vaccination rather than by infection) (24). The Hong Kong fifth wave showed similar mortality as in previous waves in unvaccinated individuals, and double risk of mortality among those unvaccinated compared to vaccinated (10). In Australia, with low levels of cases in previous waves, mortality in the Omicron wave was the highest to date, compared to the reduced mortality seen in South Africa, Israel and a number of European countries (25).

The reduced mortality in the Omicron BA.4/BA.5 wave was more specifically presumed to be related to the high prevalence of humoral and cell-mediated immunity in South Africa. Studies of humoral immunity reported SARS-CoV-2 antibody sero-prevalence estimates of 71% (26) and 73% (3) before the Omicron BA.1/BA.2 wave, while estimates after the Omicron BA.1/BA.2 wave but just before the Omicron BA.4/BA.5 wave were 97% (27) and 91% (28). The latter serosurvey indicated that 61% of individuals had serological evidence of SARS-CoV-2 infection during the Omicron BA.1/BA.2 wave in South Africa (28). In addition, 49% of adult South Africans had received at least one vaccine dose by April 2022, although the uptake of COVID-19 booster doses was low, with less than 5% of people having received a booster (29) (The South African vaccination programme is described in Supplemental Methods). The prevalence of cell-mediated following natural infection or vaccination is likely to follow a similar trend in South Africa.

Some have expressed uncertainty about ascribing lower severity in the Omicron waves to an intrinsically less virulent variant (30). The propensity of Omicron BA.1 to infect the upper rather than the lower airways suggest a mechanism for less severe disease (2) and mice studies suggested less pathogenic disease (31). However, an animal study suggested that Omicron BA.4/BA.5 in the absence of prior protection may be more pathogenic than Omicron BA.2 (32).

Admission incidence in the fifth wave was the lowest seen to date in individuals aged 5 years and older. However, in children below 5 years, where vaccination is not yet provided in South Africa, admission incidence in the BA.4/BA.5 wave is similar to the Beta wave and higher than in the D614G wave. The U-shaped distribution of COVID-19 admissions in children aged <5 years is similar to what is observed for other severe respiratory illness including respiratory infection related to influenza (33). This pattern is likely due to the immunity gap with young children having lower infection rates than adults and not being eligible for vaccination. Modellers have suggested that characteristics of the shift towards endemicity include a transition in the age structure once the disease reaches seasonal endemism (34).

A strength of this analysis is its high-quality national database and linked data on COVID-19 vaccination and prior infection for the comparisons in Omicron BA.4/BA.5 severity to previous variants. This study has several limitations. Reported cases are dependent on case ascertainment that is influenced by uptake of testing. Testing strategies for determining cases have changed over time, and testing rates have been lowest during the Omicron BA.4/BA.5 wave compared to any previous waves. However, testing of hospitalised patients presenting with suggestive symptoms has changed little and the criteria for hospital admission with COVID-19 has not changed much over time, except that lower admissions in the two Omicron waves resulted in more hospital bed capacity than in previous waves, so more patients could have been admitted with milder disease due to available bed capacity.

A further limitation is that the DATCOV database makes it difficult to discern between those admitted for COVID-19 and those admitted to hospital coincidentally with SARS-CoV-2 infection. The DATCOV dataset utilises wave period as a proxy for variants and does not contain individual-level data on infecting lineage, however each wave has correlated well with a particular VOC. While DATCOV is able to link with EVDS and contains full vaccination history of all COVID-19 admissions, the analysis did not take into account time since last vaccine dose to factor in the effect of waning immunity. Also, the South African vaccination programme eligibility changed over time during this period starting with health care workers and individuals older than 60 years, then expanding to other age groups and later introducing booster doses (detailed in the methods section). In a small number of cases, when national identification numbers are not available in either dataset being linked, linkage required “fuzzy” matching using first name, surname and date of birth, and may therefore be incomplete in correctly ascertaining prior infection and vaccination status though this would have minimal impact as most matching is on identification numbers. Finally, the data are likely to be incomplete on prior SARS-CoV-2 infections as infection and reinfection are substantially under-ascertained due to the high proportion of asymptomatic infections and challenges in testing.

## CONCLUSION

There were fewer SARS-CoV-2 cases, lower risk of hospitalisation and a substantially reduced risk of in-hospital mortality in the Omicron BA.4/BA.5 wave compared to all the previous waves, including the Omicron BA.1/BA.2 wave. This trend is different in young children, who are not yet included in South Africa’s vaccination strategy. The overall trend towards lower severity, reduced hospitalisations and fewer deaths is likely due to combination of lower viral virulence and increasing immunity, especially hybrid immunity from increasing vaccination coverage against a backdrop of widespread natural infection.

## Data Availability

The dataset analysed for the manuscript is available upon reasonable request. The data dictionary is available at request to the corresponding author

## ACKNOWLEDGEMENTS

We acknowledge the NICD team responsible for reporting test, case and hospitalisation data. Thanks to the National Department of Health and the Council for Scientific and Industrial Research for implementation support and access to vaccination rates (in particular Melissa Burgess and Lauren Hankel), the NICD for support and oversight, and the Network for Genomics Surveillance in South Africa (NGS-SA) for sequence frequencies. Our gratitude to the laboratories, clinicians and data teams in all public and private sector hospitals throughout the country reporting cases and hospitalisation data, who are acknowledged and listed as the DATCOV author group: https://www.nicd.ac.za/diseases-a-z-index/covid-19/surveillance-reports/daily-hospital-surveillance-datcov-report/

## CONTRIBUTORSHIP

WJ, LO, CM contributed to literature search. WJ, LB, CC, MM, LO and CM contributed to study design and refining methods of analysis. LO, WJ, CM and RW contributed to data analysis, and creation of tables and figures. WJ, MG, CC, RW and LO contributed to data interpretation and initial draft. WJ, RW and LO drafted the initial manuscript and all other co-authors contributed scientific inputs equally towards the interpretation of the findings and the final draft of the manuscript. WJ, LO, CM and RW have verified the underlying data.

---

**Supplementary Table S1:** Missing data and characteristics of COVID-19 hospitalised patients reported to DATCOV, 5 March 2020-28 May 2022, DATCOV, South Africa (N=212,624).

| Characteristic | Missing data<br>n (%) | Unimputed<br>n (%) | Imputed<br>% (95% CI) |
| --- | --- | --- | --- |
| **Sex** | **438 (0.2)** | **N=218,827** | **N=219,265** |
| Female |  | 121,937 (55.7) | 55.7 (55.5-55.9) |
| Male |  | 96,890 (44.3) | 45.6 (45.2-46.1) |
| **Age (in years)** | **Complete** | **N=219,265** | **N=219,265** |
| <20 |  | 8,605 (3.9) | 3.9 (3.8-4.0) |
| 20-30 |  | 44,605 (20.3) | 20.3 (20.2-20.5) |
| 40-59 |  | 86,095 (39.3) | 39.3 (39.1-39.5) |
| 60-69 |  | 41,077 (18.7) | 18.7 (18.6-18.9) |
| 70-79 |  | 24,943 (11.4) | 11.4 (11.2-11.5) |
| ≥80 |  | 13,940 (6.4) | 6.4 (6.3-6.5) |
| **Race** | **73,389 (33.5)** | **N=145,876** | **N=219,265** |
| White |  | 12,661 (8.7) | 10.9 (10.7-11.1) |
| Mixed |  | 10,053 (6.9) | 11.2 (11.0-11.4) |
| Black |  | 114,571 (78.5) | 72.2 (71.9-72.5) |
| Indian |  | 8,591 (5.9) | 5.7 (5.6-5.8) |
| **Comorbidity** | **55,915 (25.5)** | **N=163,350** | **N=219,265** |
| No |  | 102,252 (62.6) | 61.4 (61.1-61.6) |
| Yes |  | 61,098 (37.4) | 38.6 (38.4-38.9) |
| **Health sector** | **Complete** | **N=219,265** | **N=219,265** |
| Private |  | 105,409 (48.1) | 48.1 (47.9-48.3) |
| Public |  | 113,856 (51.9) | 51.9 (51.7-52.1) |
| **Province** | **Complete** | **N=219,265** | **N=219,265** |
| Eastern Cape |  | 28,671 (13.1) | 13.1 (12.9-13.2) |
| Free State |  | 11,759 (5.4) | 5.4 (5.3-5.5) |
| Gauteng |  | 59,548 (27.2) | 27.2 (27.0-27.3) |
| KwaZulu-Natal |  | 42,712 (19.5) | 19.5 (19.3-19.6) |
| Limpopo |  | 8,110 (3.7) | 3.7 (3.6-3.8) |
| Mpumalanga |  | 8,287 (3.8) | 3.8 (3.7-3.9) |
| North West |  | 11,343 (5.2) | 5.2 (5.1-5.3) |
| Northern Cape |  | 3,494 (1.6) | 1.6 (1.5-1.6) |
| Western Cape |  | 45,341 (20.7) | 20.7 (20.5-20.8) |
| **Wave period** | **51 (<0.1)** | **N=219,214** | **N=219,265** |
| Delta |  | 400 (0.2) | 0.2 (0.2-1.2) |
| Omicron BA.1 |  | 1,449 (0.7) | 0.7 (0.6-0.7) |
| Omicron BA.4/BA.5 |  | 5,787 (2.6) | 2.6 (2.6-2.7) |
| **Vaccination** | **Complete** | **N=219,265** | **N=219,265** |
| No |  |  |  |
| Yes |  |  |  |
| **Prior infection** | **Complete** | **N=219,265** | **N=219,265** |
| No |  |  |  |
| Yes |  |  |  |
| **In-hospital outcome** | **Complete** | **N=219,265** | **N=219,265** |
| Discharged alive |  | 168,228 (76.7) | 76.7 (76.5-76.9) |
| Died in hospital |  | 51,037 (23.3) | 23.3 (23.1-23.5) |

**Supplementary Figure S1.**
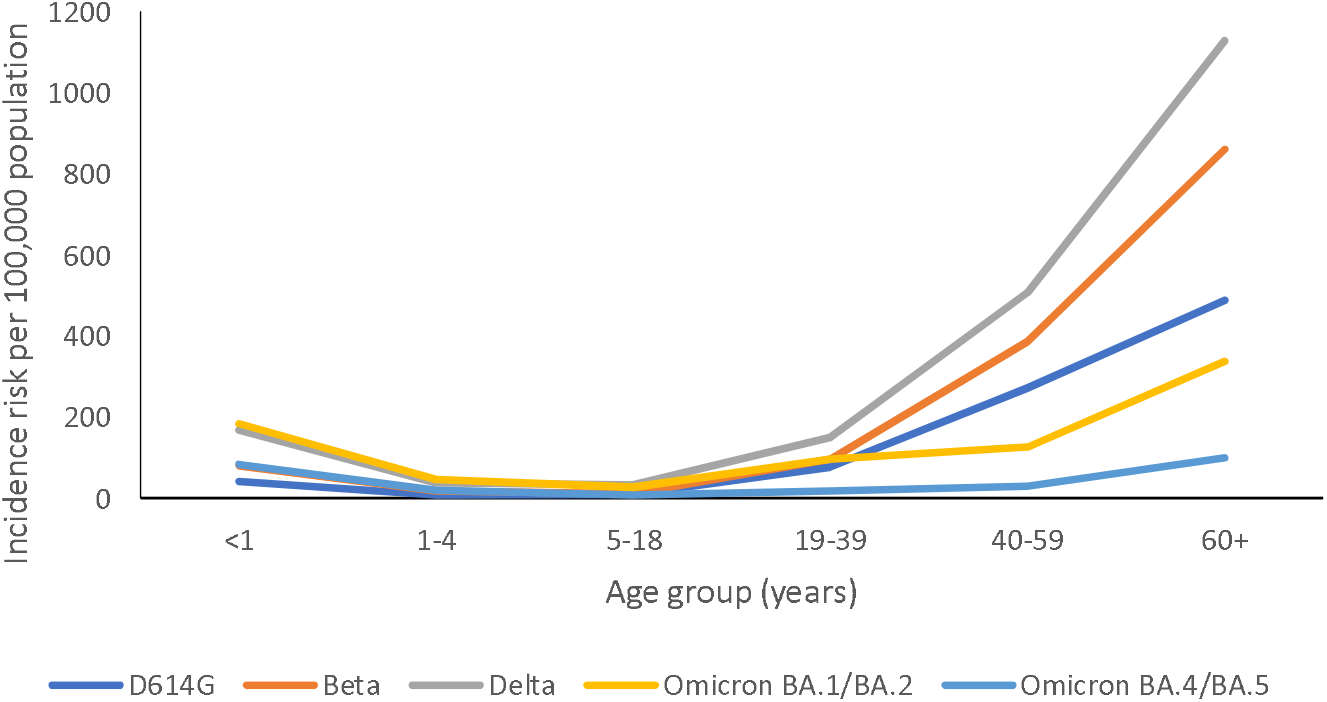
COVID-19 admissions incidence risk per 100,000 population, by age group (in years) and wave period, South Africa, 5 March 2020-28 May 2022 (N=438,965)

